# Efficacy versus abundancy: which vaccination schemes can better prohibit deaths and active infections?

**DOI:** 10.1101/2021.09.02.21263041

**Authors:** Omar El Deeb, Maya Jalloul

## Abstract

In this paper, we introduce a general novel compartmental model accounting for the effects of vaccine efficacy, deployment rates and timing of initiation of deployment. It consists of compartments corresponding to susceptible, vaccinated susceptible, infectious, vaccinated infectious, active, and dead populations with various vaccine efficacies and vaccination deployment rates.

We simulate different scenarios and initial conditions, and we find that the abundance and higher rate of deployment of low efficacy vaccines would lower the cumulative number of deaths in comparison to slower deployment of high efficacy vaccines. However, the latter can lower the number of active cases and achieve faster and higher herd immunity. We also forecast that, at the same daily deployment rate, the earlier introduction of vaccination schemes with lower efficacy would also lower the number of deaths with respect to a delayed introduction of high efficacy vaccines, which can however, still achieve lower numbers of infections and better herd immunity.

## 1 Introduction

Humanity has been struggling with viral pandemics and infectious diseases that caused great catastrophes throughout the entire history of mankind. Since the Athenian, Antonine and Justinian Plagues to the Black Death, Spanish Flu, Cholera, and Smallpox up until the HIV pandemic, SARS, Swine flu, Ebola outbreak and, most recently, the Coronavirus infection, pandemics have been a major source of disease, death, economic crises and political turmoils [1, 2, 3]. However, those outbreaks also lead to major discoveries and advancements in sciences and public health, especially in medicine, pharmaceuticals, vaccines and development of public policies [4, 5, 6].

The Coronavirus disease (COVID-19) associated to the severe acute respiratory syndrome coronavirus 2 (SARS-CoV-2) continues as a main cause of hospitalization and death and as a main public health risk since the first case was registered in Wuhan, China in December 2019 [7]. It was declared as a global pandemic on March 11, 2020 [8] after spreading from China into several Asian and European countries and consequently into tens of countries across the world. By August 23, 2021, the world has suffered about 213 million registered infections and 4.45 million deaths associated to COVID-19 in 222 different countries, territories and entities [9]. Health systems were put under enormous pressure, and the consequent mitigation measures and associated closures and lockdowns taken across the world for lengthy periods of time contributed to a global economic slowdown and recession in several countries [10, 11], in addition to a heavy negative impact on education, employment, tourism and social activities [12, 13, 14] exacerbating existing political discontent and stirring political unrest [15].

Similarly to the other respiratory pathogens, airborne transmission of SARS-CoV-2 occurs by inhaling droplets loaded with the virus emitted by infectious people. Infection can also occur through mucous membranes like the eyes, nose or mouth [16].

Many variants of the virus emerged with an increased risk to global public health. The World Health Organization (WHO) characterized variants of interest (VOI) and variants of concern (VOC) to track and monitor [17]. The main variants of concern have stronger capabilities of trasmissibility and health risks, and they are labeled by the WHO as the Alpha, Beta, Gamma and Delta variants which were first documented in the United Kingdom, South Africa, Brazil and India respectively. The delta variant is the most dangerous mutation yet as it is the most transmissible form of the virus, and relatively deadlier too [18]. The recent emergence of variants creates a major cause of concern since they can lead to an epidemic rebound especially with the possibility of vaccine resisting and the emergence of deadlier and/or more transmissible future variants. Increased viral transmission means higher probability of emergence of variants as well [19].

Several types of vaccines were developed and approved by global (WHO) or by national health agencies, using different techniques like the viral vector vaccines (Oxford/AstraZeneca, Sputnik V/Gamaleya, Janssen), genetic vaccines (Pfizer/BioNTech, Moderna), inactivated vaccines (Sinovac, Sinopharm, Bharat) and protein vaccines (Novavax, Sanofi) [20]. By August 23, 2021, a total of about 4.97 billion doses of those vaccines were administered with main concentration in China, USA, India and Europe [21]. Other vaccines, inhaled aerosol dry vaccines and antiviral drugs are under extensive research and development [22, 23, 24]. Antibody therapies are under development as well and they offer an effective treatment for the most seriously ill patients, but they remain expensive and in short supply [25].

A global roll-out of vaccines is needed to guarantee a swift elimination of the infection, and for this end vaccines need to be available, affordable, and accessible at a global scale. In high-income countries, large stocks were produced and purchased, combined with active logistic and public health resources, but in low-income countries, vaccination is still slow mainly due to insufficiency of vaccines. The WHO has called for more equity and stronger support for its COVAX initiative in order to supply wider vaccine access for poor countries, but statistics show that the gap remains deep as many rich countries have already bought multiple times of doses needed per person [26]. A similar gap also arises on the national level as health illiteracy, religious beliefs and sometimes political partisanship are slowing down vaccinations and creating irregular immunity rates among different regions or groups [27]. As such, the global efforts to stop the spread remains far from successful, and the priority should be focused to optimize the use, deployment and timing of available vaccines, whether those recognized by the WHO or by national health agencies across the globe in order to prevent further deaths as well as emergence of new variants.

The spread patterns of infectious diseases have been and are being actively studied and simulated using several mathematical and computational models. A wide variety of techniques were employed ranging from compartmental models, Agent-Based models (ABM), spatio-temporal analysis to data-driven analysis and artificial intelligence [28, 29]. The first compartmental model was the famous Kermack–McKendrick Susceptible-Infectious-Removed (SIR) model which divides the total population under study into three compartments where agents would move from one compartment to another upon infection or removal through recovery or death [30]. Numerous variations of this model were introduced to account for characteristics and dynamics of different diseases as well as in simulating similar interactions and spreads in the social and behavioral sciences [31, 32, 33]. The research on COVID-19 spread extensively employed SIR models that were improved to account for other effects like exposure, travel, quarantine, vaccination, public measures and many other alterations to best describe the underlying features of the contagion [34, 35, 36, 37, 38], as well as general and country specific ABMs, spatio-temporal and data driven studies [39, 40, 41, 42, 43].

However, with the mass introduction of various globally and nationally recognized vaccines with varying efficacies, and with the varying degrees of availability and logistic capabilities across the globe, an important question arises for decision makers: which vaccine to choose to deploy in a certain country from the available options in that country, given their efficacies, their availability timeline and expected deployment rate of each. This is a primary issue for public health officials aiming at minimizing the number of deaths and/or the number of active infections, that was not thoroughly discussed and analyzed in the literature. This paper fills the gap by studying the trade-off between vaccine efficacy and abundance and then between efficacy and time of availability, and the corresponding expected outcomes for deaths and infections.

The paper is organized as follows: section 2 presents the novel theoretical compartmental model, section 3 puts forward the results and discussions about the efficacy, deployment and timing rates while the conclusion is presented in section 4.

## 2 Theoretical Model

We introduce a novel compartmental model to account for the effect of vaccines, taking into account different vaccination deployment rates (*v*), efficacies (*e*) under several scenarios of infection spread rates represented by the reproductive number (*R*_*t*_), starting with different initial conditions in relation to different numbers of infections, deaths and immune populations in different countries. The *SS*_*V*_ *II*_*V*_ *RD* model consists of 6 compartments: The susceptible unvaccinated population *S*, the susceptible vaccinated population *S*_*V*_ who may catch the virus after being vaccinated during the assumed effective immunity period depending on the efficacy of the vaccine, the infectious unvaccinated population *I*, the infectious vaccinated population *I*_*V*_ who already caught the infection after being vaccinated, the recovered population *R* who are either fully protected by the vaccine during its effective protection time range or who have already recovered from the infection and still possess immunity, and finally the dead *D*. Each population is normalized with respect to the total population so that *N* = *S* + *S*_*V*_ + *I* + *I*_*V*_ + *R* + *D* = 1.

The 6 compartments are interlinked, and their dynamics can be modeled by a system of coupled linear ordinary differential equations given by:

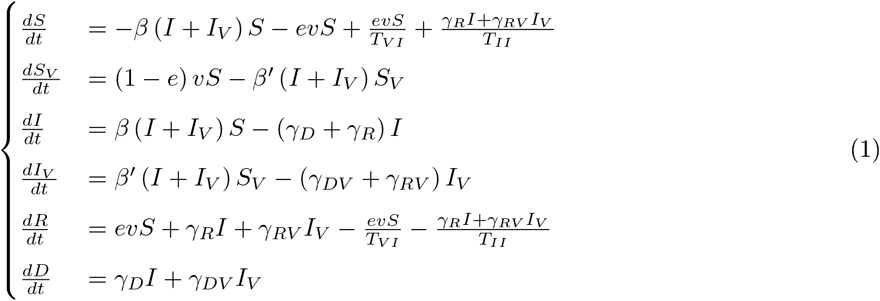

The parameter *β* is the rate of meeting between susceptible (*S*) and all infectious people (*I* + *I*_*V*_) and it can be determined from the reproductive number of the infectious spread *R*_*t*_ and the average rate of recovery of unvaccinated people*γ*_*R*_. In this sense, *β* = *γ*_*R*_*R*_*t*_. The rate of recovery of vaccinated people is given by *γ*_*RV*_ and the rate of meeting between vaccinated susceptible people with infectious people is given by *β′* = *γ*_*RV*_ *R*_*t*_. The rate of death of unvaccinated people is *γ*_*D*_ and that of vaccinated ones is *γ*_*DV*_. Finally, *T*_*VI*_ represents the average time duration of acquired vaccine immunity and *T*_*II*_ represents the average duration of acquired post infection immunity. The change in the total population is 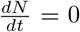, assuming that *N* remains constant during that period, neglecting other changes due to natural growth, immigration, etc…

The transfer dynamics between different compartments can be summarized according to the following:

- The susceptible *S* may become infected *I* upon contact with the infectious (vaccinated or non-vaccinated) are rate *β*. They may also become recovered *R* upon receiving a vaccine with efficacy *e* given at a daily rate *v*. In addition, recovered *R* people would become susceptible again after some time of recovery. Mainly, recovered people due to infection would become susceptible again in an infection immunity period *T*_*II*_ and recovered people due to vaccination would become susceptible again in a vaccine immunity period *T*_*VI*_.
- The vaccinated susceptible population *S*_*V*_ are the susceptibles *S* who took the vaccine but are still susceptible to infection, and they may become infectious *I*_*V*_ upon a meeting rate *β′* with other infectious people.
- The infectious compartment *I* is populated by unvaccinated susceptibles *S* bumping into other infectious agents at a rate *β*, while exit from this compartment is attributed to deaths at rate *γ*_*D*_ into compartment *D* and recoveries at rate *γ*_*R*_ into compartment *R*.
- The vaccinated infectious compartment *I*_*V*_ is formed by vaccinated susceptibles *S*_*V*_ catching the disease at a rate *β′* and diminished by people dying at rate *γ*_*DV*_ into *D* and people recovering a rate *γ*_*RV*_ into *R*.
- The recovered population *R* is formed by effectively vaccinated susceptible people coming from *S* and people surviving the infection at rates *γ*_*R*_ and *γ*_*RV*_ from infectious and vaccinated infectious populations *I* and *I*_*V*_. Simultaneously, recovered people would eventually lose their acquired immunity on average periods of *T*_*II*_ after infection and *T*_*VI*_ after vaccination, thus would exit the recovered compartment *R* back to the susceptibles *S*.
- Finally, the dead *D* increase at death rates *γ*_*D*_ and *γ*_*DV*_ among infectious and vaccinated infectious populations *I* and *I*_*V*_.

The vaccination deployment rate *v* depends on the available supply of the vaccine as well as the logistical capability and the popular demand at a given time [44]. It is taken in this model as the daily vaccination percentage of the susceptible population, with different scenarios representing slow, moderate and fast deployment rates. The vaccine efficacy varies among different employed vaccines as well as in relation to new emerging variants in addition to the possibility of supplying a single dose of a double dosed vaccine under short supply. The model accounts for a range of scenarios with low, moderate and high efficacy corresponding to the former cases. We also inspect those scenarios under different reproductive rates *R*_*t*_ which depend on different values of mitigation measures related to social distancing, protective masks, sanitation, and other factors that alter the rate of infection spread. We assume different scenarios of low, moderate, high and alternating (where *R*_*t*_ varies in a periodic pattern between high and low extrema) reproductive rates.

The numerical values of these parameters are taken in relation to available data and research. The efficacy values vary between 0.5 ≤ *e* ≤ 0.95 depending on the available vaccines [45, 46, 47, 48, 49, 50, 51]. The rate of recovery for vaccinated people 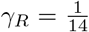 in relation of an average of 14 days needed for recovery [52], while the death rate amounts to around 2% [9] of the infected which leads to 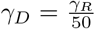. We also assume that those who are infected after vaccination would need a similar time of recovery, despite the fact that their symptoms would be much reduced [53, 54], thus *γ*_*RV*_ = *γ*_*R*_ but their death rate, as studies reveal, would be considerably lower by 70 −85% [54, 55], which is modeled through 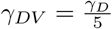. We take *T*_*VI*_ = 90 days and *T*_*II*_ = 360 days to represent expected periods of acquired immunity after recovery and after vaccination respectively [56]. The deployment rate *v* is assigned hypothetical values varying between 0.1% of the susceptible population per day for slow vaccination rollout and 1.5% for the highest pace of vaccination rollout. The reproduction rate *R*_*t*_ is assigned values of 0.7, 1.1 and 1.5 corresponding to low, medium and high reproduction rates, in addition to a fourth scenario where *R*_*t*_ alternates sinusoidally between 0.4 and 2 according to the relation 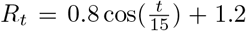 where *t* represents the time in days, with a period of around 94 days to simulate the effect of the consecutive waves of the spread of the infection, as observed empirically [57, 58].

We simulate this model under different combinations of efficacy *e*, deployment rate *v* and reproductive number *R*_*t*_ to analyze the corresponding cumulative numbers of infected, recovered and dead populations.

## 3 Results and discussions

### 3.1 Efficacy vs abundance

We simulate the theoretical model introduced in (1) to determine relative numbers of active cases, infected vaccinated cases, immune population and total cumulative deaths under nine different combinations of efficacies (*e*) of available vaccines and their deployment rates (*v*) given by: *e* = 92%, *v* = 0.5%, *e* = 92%, *v* = 0.3%, *e* = 92%, *v* = 0.1%, *e* = 72%, *v* = 0.7%, *e* = 72%, *v* = 0.5%, *e* = 72%, *v* = 0.3%, *e* = 55%, *v* = 1.5%, *e* = 55%, *v* = 1% and *e* = 55%, *v* = 0.7% in order to compare the levels of infection, death and immunity between higher efficacy vaccines at lower abundance and lower efficacy vaccines with more abundance or deployment rate, together with middle values between them. We also account for different possible current situations of infection spread that may differ from one country to another by taking two hypothetical initial conditions corresponding to countries which already achieved high levels of vaccination (40.6% vaccinated population 1.8% currently infected and 0.18% dead) and countries with low current levels of vaccination (3.1% vaccinated population 0.46% currently infected and 0.11% dead). In this sense, the results of this forecast are not country specific but are of global significance. We repeat this simulation for different levels of infection spread modeled through low, medium, high and alternating reproductive rates *R*_*t*_ defined before. The results are displayed in figures 2,3,4 and 5 respectively.

**Figure 1:**
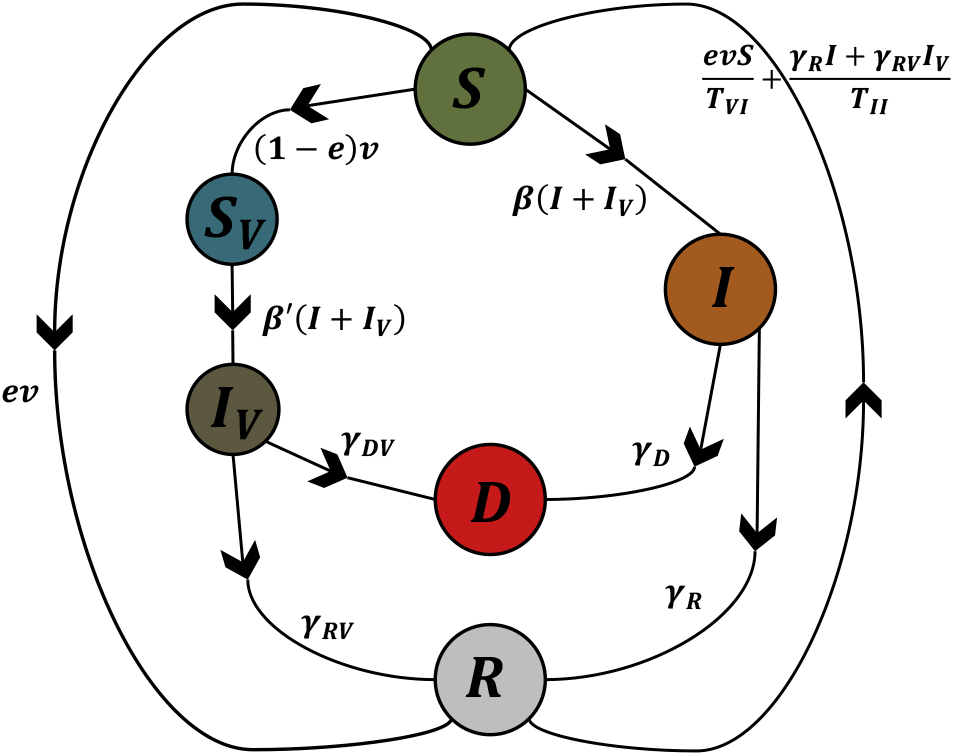
A schematic diagram of the *SS*_*V*_ *II*_*V*_ *RD* model showing the six compartments of the model (Susceptible, Vaccinated susceptible, Infectious, Vaccinated Infectious, Recovered and Dead) together with their transfer dynamics and the coefficients relating transfers among different compartments.

**Figure 2:**
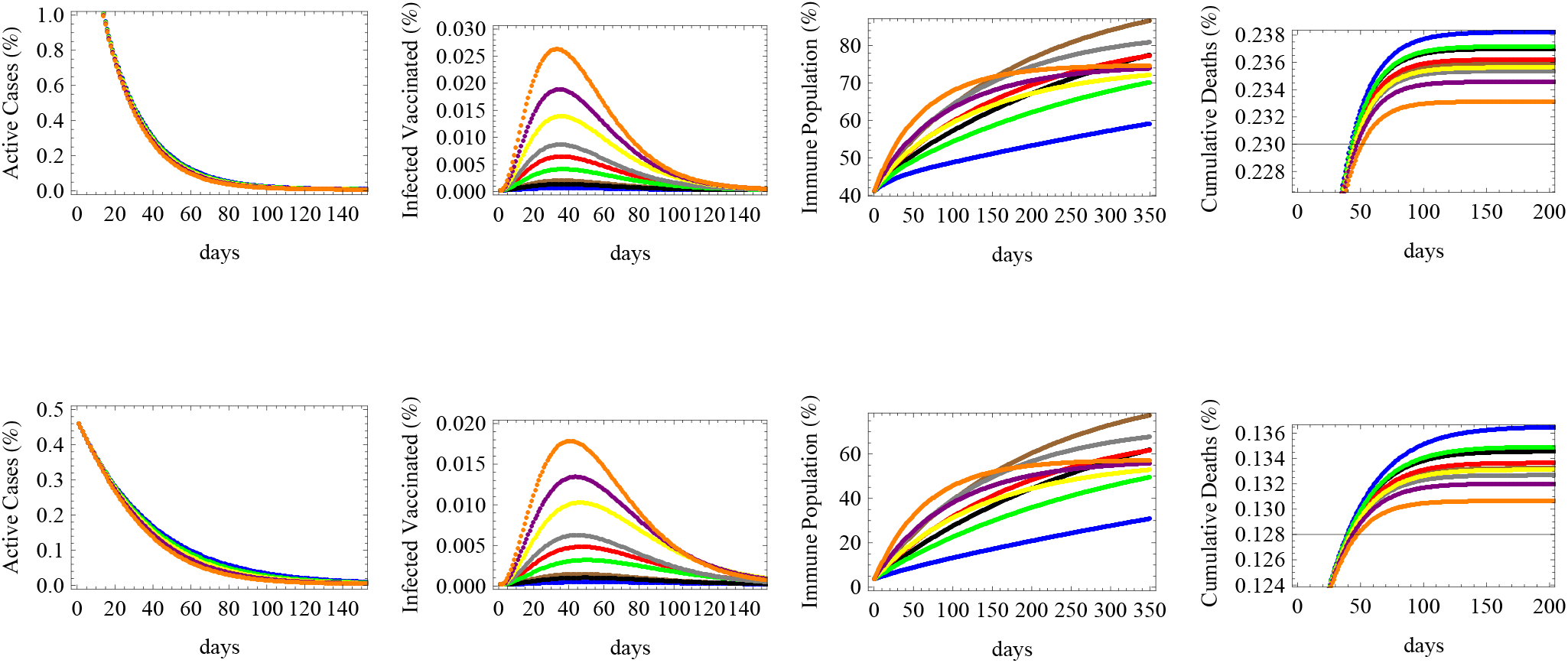
The relative numbers of active cases, infected vaccinated cases, immune population and total cumulative deaths are shown for a low reproductive rate *R* = 0.7 under initial conditions of: 40.6% vaccinated population 1.8% currently infected and 0.18% dead (upper row) and 3.1% vaccinated population 0.46% currently infected and 0.11% dead (lower row) for nine different vaccination scenarios. The forecast corresponds to vaccine efficacy (*e*) and daily deployment rate (*v*) of: *e* = 92%, *v* = 0.5% (in brown), *e* = 92%, *v* = 0.3% (in black), *e* = 92%, *v* = 0.1% (in blue), *e* = 72%, *v* = 0.7% (in gray), *e* = 72%, *v* = 0.5% (in red), *e* = 72%, *v* = 0.3% (in green), *e* = 55%, *v* = 1.5% (in orange), *e* = 55%, *v* = 1% (in purple) and *e* = 55%, *v* = 0.7% (in yellow).

**Figure 3:**
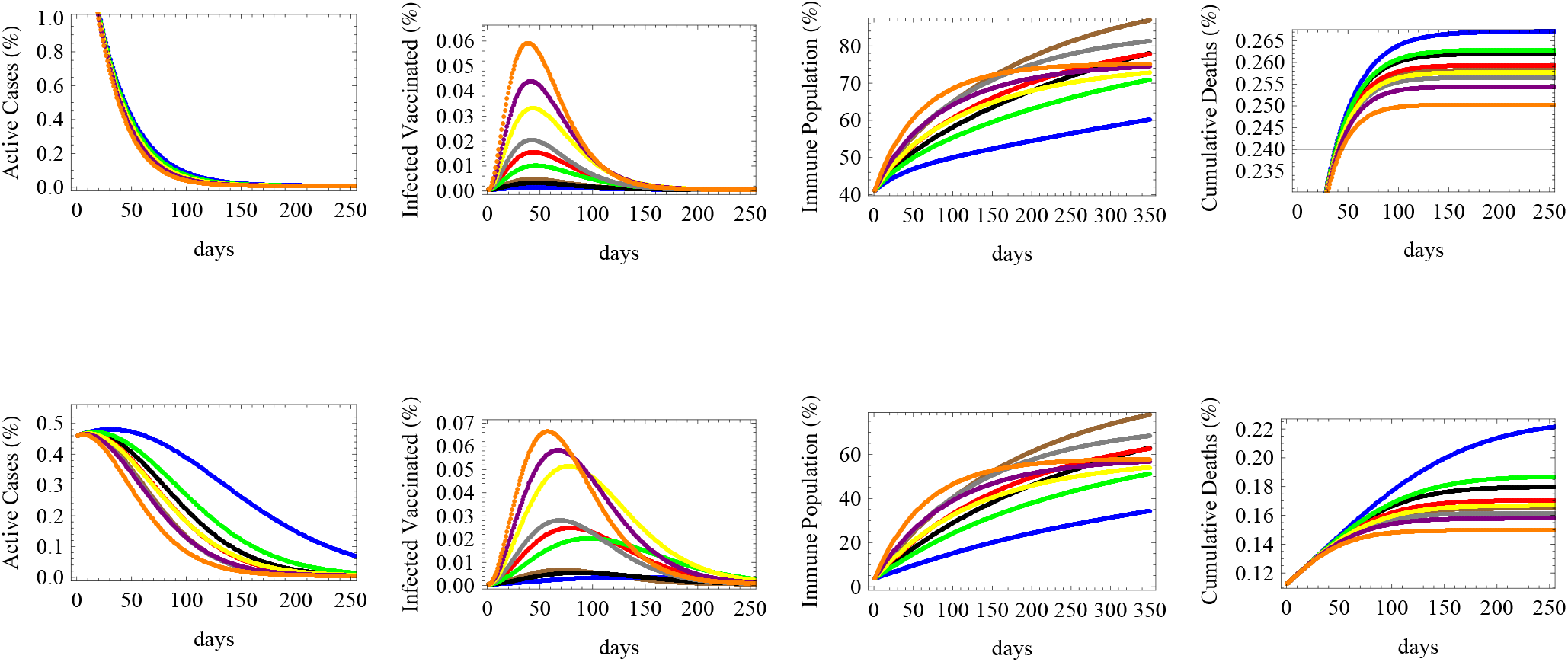
The figure shows the relative numbers of active cases, infected vaccinated cases, immune population and total cumulative deaths for a medium reproductive rate *R* = 1.1 with the same two initial conditions and nine vaccination scenarios of figure 2.

**Figure 4:**
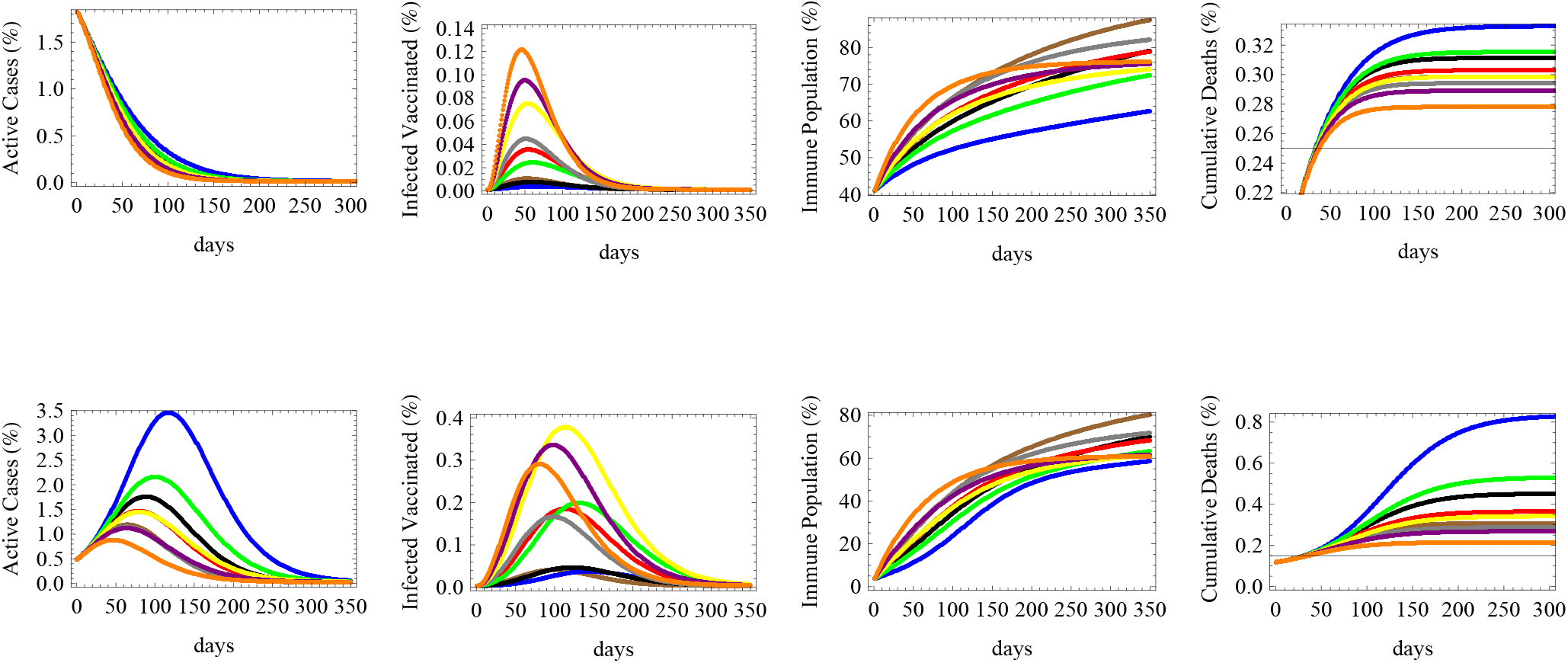
The figure shows the relative numbers of active cases, infected vaccinated cases, immune population and total cumulative deaths for a high reproductive rate *R* = 1.5 with the same two initial conditions and nine vaccination scenarios of figure 2.

**Figure 5:**
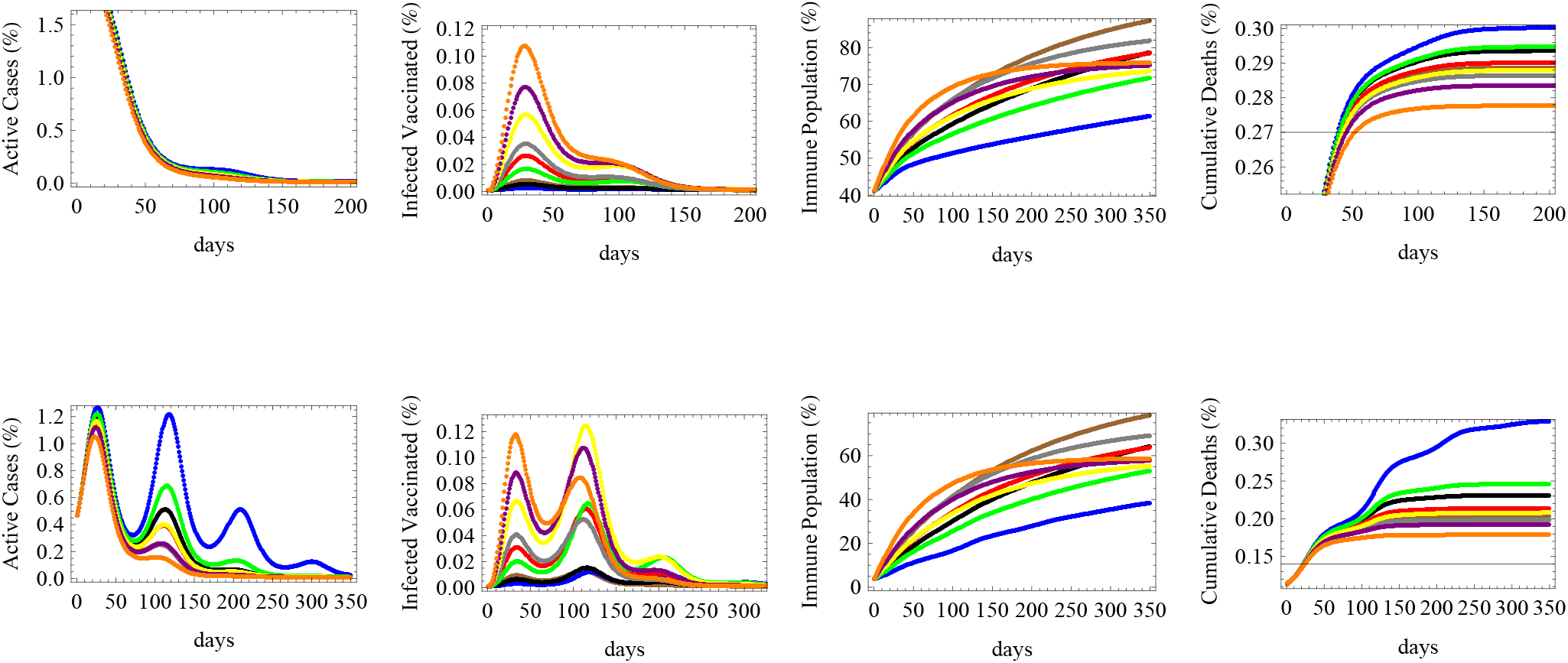
The figure shows the relative numbers of active cases, infected vaccinated cases, immune population and total cumulative deaths for an alternating reproductive rate, varying sinusoidally between *R* = 0.4 and *R* = 2, with the same two initial conditions and nine vaccination scenarios of figure 2.

Figure (2) shows that when the reproductive rate is low (*R*_*t*_ = 0.7), the number of active cases would fall down quickly under all vaccination schemes, for various efficacies and deployment rate, both in countries that are in the middle or early stages of the spread (upper and lower figures respectively). The number of infected vaccinated people is the highest for the low efficacy vaccine. Herd immunity would be achieved optimally under fast deployment of the high efficacy vaccine and the middle efficacy vaccine, while the lowest immunity is achieved under the slowest deployment of the high efficacy vaccine. This shows that the deployment rate is an essential factor in combination with efficacy for achieving immunity. The figure also shows that the number of deaths is most reduced under the two fastest deployment schemes of the low efficacy vaccine, followed by the fastest deployment rate of that of middle efficacy. The latter result shows that to reduce deaths, the most essential measure is to ensure the the availability and the fast deployment of the vaccine, while reaching herd immunity and decreasing infections (thus lowering the pressure on the health sector and decreasing the possibility of appearance of new mutations) depends relatively more on its efficacy.

In the case of a medium reproductive rate (*R*_*t*_ = 1.1) corresponding to a slow increase in the number of infections in absence of vaccine, figure (3) shows that the number of active cases would decrease in countries that are in their middle stages of infection, while they would keep on increasing for a limited interval of time in countries that are in their early stages of spread. The duration needed for the active cases to start decreasing is shortest for the vaccine with fastest deployment rate (despite its low efficacy), while that with the slowest deployment rate (with high efficacy) needs more time to decrease the number of active cases. The highest number of infected vaccinated people would be obtained through the rapidly deployed low efficacy for countries in their middle stages of spread, while for early stage countries, each scenario of deployment of the low efficacy vaccine would lead to maximal infections under different times. In a similar manner to what happens for low reproductive rate, immunity will be maximally obtained under the deployment of high efficacy vaccines at high rates, while the least deaths are achieved under the fastest deployment rates of vaccines.

For a country under a fast spread scenario simulated by *R*_*t*_ = 1.5, figure (4) shows that the number of active cases would fall in a middle spread stage country, though it takes more time than that with lower *R*_*t*_. However, in early spread countries, the number of active cases will rise and peak after a period of time before falling down after a relatively long time (around 100 − 200 days) of vaccination. The slowest vaccination schemes of the low and middle efficacy vaccines would reach the highest peaks and need more time for controlling the spread while the fastest vaccination schemes would achieve it the quickest. The number of infected vaccinated people would be the largest in the case of the low efficacy vaccine deployed rapidly in a middle spread stage country, while for a country at early spread stages, the two slowest deployments of the low efficacy vaccine lead to the highest infections among the vaccinated population. Immunity will be maximally obtained under the deployment of high efficacy vaccines at high rates, while the least deaths are achieved under the fastest deployment rates of vaccines, like what was shown before for low and medium spread rates, but with a much higher magnitude of deaths in all scenarios due to a higher reproductive rate.

In the more realistic case of an alternating reproductive number varying between 0.4 ≤ *R*_*t*_ ≤ 2 and corresponding to consecutive patterns of low and high spread waves, we realize that in a country at a middle spread stage, all vaccination strategies would lead to bringing down the number of active cases with a small peak arising after the return of the next wave, while in a country at early stages of spread, the slow deployment of vaccines, whether with high or low efficacies, would cause the re-emergence of several peaks of infections and high number of active cases. Only the fast deployment of vaccines of various efficacies would lead to curb the number of infections just after the first peak of spread. The number of infected vaccinated people would be the highest for the case of fast deployment of low efficacy vaccines in countries with middle stages of spread while stronger peaks of infections among the vaccinated people would occur under slower vaccination scenarios of low efficacy vaccine during the second wave in countries with early spread stages. The highest herd immunity levels would be attained under the scenarios of fastest deployments of high and medium efficacy vaccines in both categories of countries under early or middle stages of spread after 3 − 4 months, despite the early lead and the sharp rise in the immune population during the first few weeks of fast deployment of low efficacy vaccines. The cumulative number of deaths would rise in jumps corresponding to successive waves of spread especially in early stage countries, but in both cases, the fastest vaccination schemes of low efficacy vaccines would ultimately lead to the minimal number of deaths while the slowest pattern of high efficacy vaccine deployment would result in the highest cumulative rate of deaths. Those simulations are displayed in figure (5).

### 3.2 Efficacy vs time of availability

An important aspect of vaccination strategies that we consider in this paper is the question of vaccine efficacy versus time of availability or start of deployment. In this simulation, we forecast the relative number of active cases, infected vaccinated people, the immune population and the cumulative number of deaths under low, medium and high reproductive rates *R* = 0.7, 1.1 and 1.5 respectively in figures (6, 7 and 8). Four cases simulating the effect of efficacy and the time of initiation of the vaccination process were considered: a vaccine of low efficacy *e* = 52% to start deployment immediately (red), a vaccine of medium efficacy *e* = 72% to start deployment immediately (black), a medium efficacy vaccine with *e* = 72% to start deployment after 30 days (blue) and a vaccine of high efficacy *e* = 92% to start deployment after 30 days (brown), all being deployed at an equal rate of the susceptible population per day. As in previous simulations, the upper row corresponds to countries at middle stages of spread with initial conditions of 40.6% vaccinated population 1.8% currently infected and 0.18% dead while the lower row corresponds to countries at early stages with 3.1% vaccinated population 0.46% currently infected and 0.11% dead.

**Figure 6:**
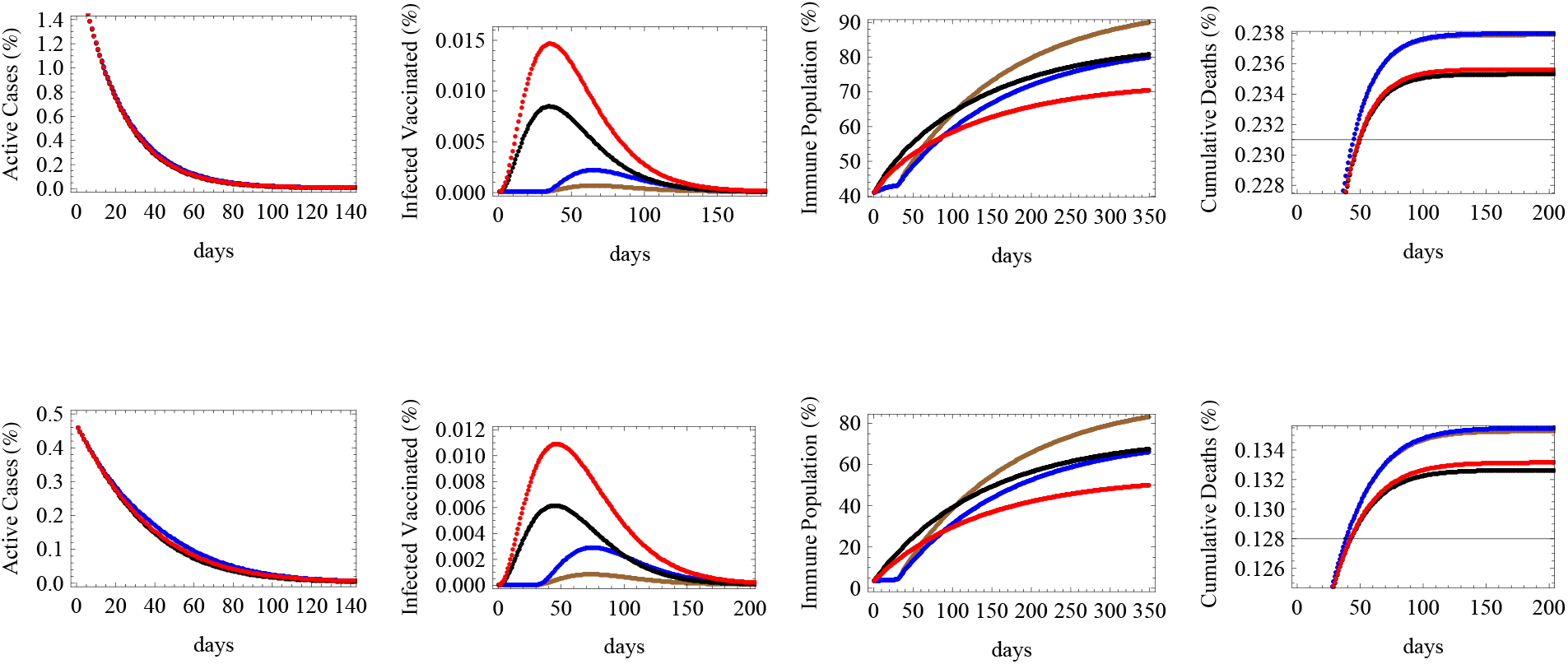
The figure shows the expected relative numbers of active cases, infected vaccinated cases, immune population and cumulative deaths at a low reproductive rate *R* = 0.7 in relation to four cases simulating the effect of efficacy and the time of initiation of the vaccination process: a vaccine of efficacy *e* = 52% to start deployment immediately (red), a vaccine of efficacy *e* = 72% to start deployment immediately (black), a vaccine of efficacy *e* = 72% to start deployment after 30 days (blue) and a vaccine of efficacy *e* = 92% to start deployment after 30 days (brown), all being deployed at an equal rate of 0.7% of the susceptible population per day. The upper row corresponds to initial conditions of 40.6% vaccinated population 1.8% currently infected and 0.18% dead while the lower row corresponds to 3.1% vaccinated population 0.46% currently infected and 0.11% dead.

**Figure 7:**
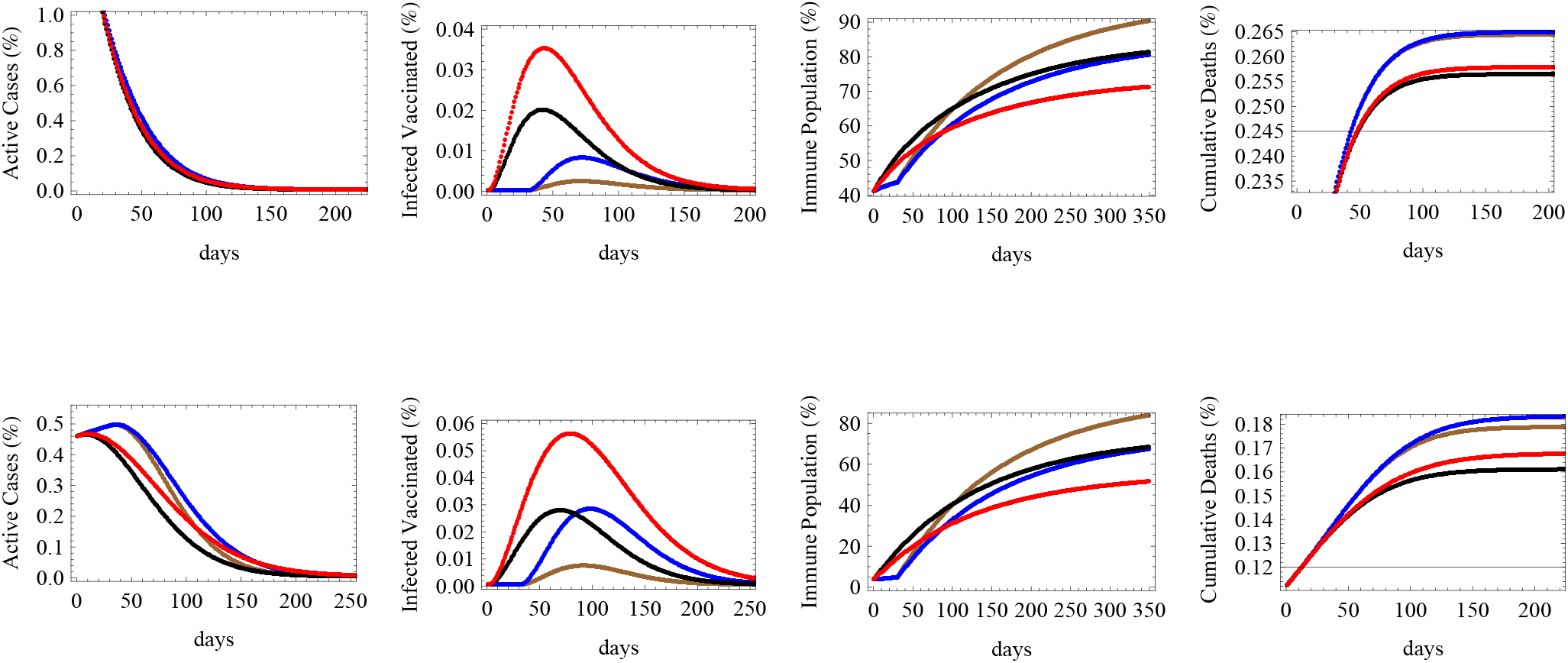
The figure shows the expected relative numbers of active cases, infected vaccinated cases, immune population and cumulative deaths corresponding to a medium reproductive rate *R* = 1.1 with same initial conditions, efficacies and time of initiation as those detailed in figure 6.

**Figure 8:**
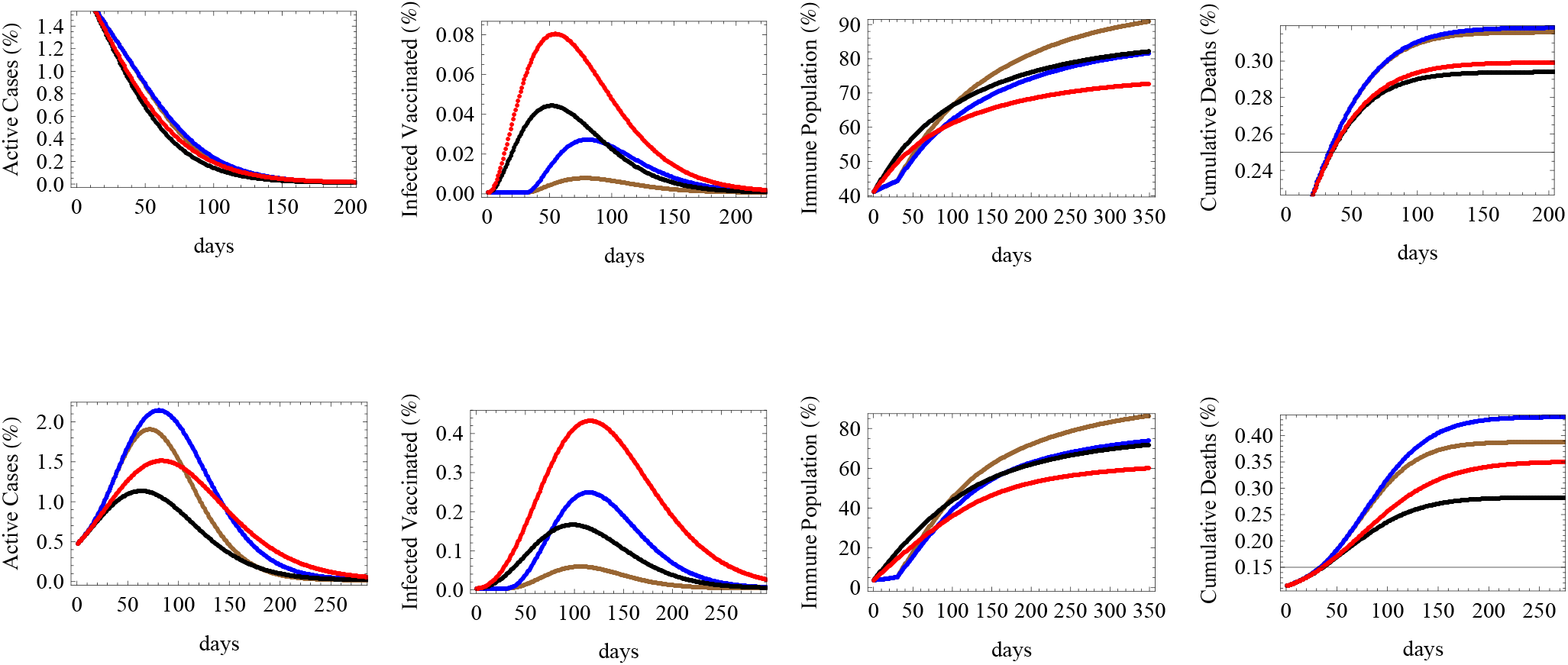
The figure shows the expected relative numbers of active cases, infected vaccinated cases, immune population and cumulative deaths corresponding to a high reproductive rate *R* = 1.5 with same initial conditions, efficacies and time of initiation as those detailed in figure 6.

Under the circumstances of low reproductive rate *R*_*t*_ = 0.7 depicted in figure (6), it is clear that the number of active cases will fall significantly under the four efficacy-timing schemes in both early and middle spread stage categories. The number of infected vaccinated people in both categories would occur under the scenario of immediate deployment of low efficacy vaccines. Herd immunity would be maximally attained through the adoption of high efficacy vaccines being deployed with a one month period of delay while it would be the lowest using low efficacy vaccines deployed immediately. On the level of cumulative deaths, the least number of deaths is realized under the scenario of immediate deployment of medium then low efficacy vaccines, while the delay would raise the number of deaths even while using high efficacy vaccines.

Figure (7) simulates same efficacy-timing scenarios under medium reproductive rate *R*_*t*_ = 1.1. We notice that in countries in their middle stage of infection spread, the number of active cases would fall quickly under all scenarios, while in early stage countries, the late deployment scenarios would cause a rise and a peak in infections before falling down significantly, whereas the quick deployment of low or medium efficacy vaccines would lower the active cases faster during the first few months. We also notice that in about 100 days, the number of active cases due to a delayed high efficacy vaccine will catch up and fall below the expected active cases under an immediately deployed low efficacy vaccine. The number of infected vaccinated people is the highest for the lowest efficacy vaccine and the lowest for the highest efficacy vaccine in both country categories. Similarly, for both categories, herd immunity is maximally attained using the delayed high efficacy vaccine rather than the immediate low efficacy one which provides the lowest percentile of immune population. However, regarding deaths, the lowest numbers of deaths are attributed to the immediate deployment of medium then low efficacy vaccines, while a delayed deployment will cause more deaths even while using high and medium efficacy vaccines.

Under the scenario of high reproductive number simulated by *R*_*t*_ = 1.5, the number of active cases would fall slowly in countries that are in their middle stages of vaccination and spread, while it would rise and peak under all vaccination scenarios in early stage countries. The highest peaks are attributed to delayed vaccinations even though infections would fall rapidly once vaccination starts, and eventually the corresponding number of active cases would fall below that of low efficacy vaccine being deployed immediately. Immediate deployment also helps to flatten the curve on infections, thus reducing the expected peak number of cases. The maximum number of infected vaccinated people would correspond low efficacy vaccine deployed immediately while the lowest corresponds to the high efficacy vaccine deployed late, for both country categories. Similarly, herd immunity is maximally achieved by the high efficacy vaccine despite being introduced late while the lowest level of immunity is caused by the low efficacy vaccine despite early introduction. The immediate deployment of medium efficacy vaccine minimizes the number of deaths, followed by the immediate low efficacy vaccine, while late deployment raises the number of deaths for both country categories, but with higher relative differences among the death outcomes for middle stage countries.

### 3.3 Limitations

This study takes into account all parameters related to vaccine efficacy and deployment rates under several infectious rates and initial conditions. However, there is a limiting factor related to the maximal proportion of the population who are willing or at least who would eventually take the vaccine. Here, we took this into account indirectly by using a daily deployment rate *v* which is proportional to the susceptibles *S*, and not to the total population *N*. In this sense, in the initial phases of vaccination, the number of people taking the vaccine would be the highest, but as time progresses, the number of susceptibles decreases, hence the daily number of vaccinated people decreases. It is natural to assume that, as when a country reaches a high level of vaccination, less people will be willing to get the vaccine. If vaccination rates were only connected to abundance or logistic infrastructure, they would have been linked to the total population *N*.

Due to various reasons ranging from religious and political beliefs, into non-scientific and anti-vaxxer fears, there might be a sizable sector of the society who would refuse to get vaccinated [27]. Vaccine hesitancy is not directly simulated in the model, but it is indirectly represented through relating the daily deployment rate to the number of susceptibles hence it decreases as the number of vaccinated people increases. In our scenario, immunity of this portion of the population would still be achieved through infection rather than vaccination.

## 4 Conclusion

In this paper we introduced a general novel compartmental model accounting for the vaccinated population, infected vaccinated population, active infections, and deaths with various vaccine efficacies and vaccination deployment rates.

We simulated different scenarios and initial conditions, and we showed that abundance and higher rate of deployment of low efficacy vaccines would lower the cumulative number of deaths in comparison to slower deployment of high efficacy vaccines. However, the high efficacy vaccines can better lower the number of active cases and achieve faster and higher herd immunity.

We also discovered that at the same daily deployment rate, the earlier introduction of vaccines with lower efficacy would also lower the number of deaths with respect to a delayed introduction of high efficacy vaccines, which can, however, lower the number of infections and attain higher levels of herd immunity.

## Data Availability

Data available upon request.

## References

[1] Huremovic, D., Brief History of Pandemics (Pandemics Throughout History). Psychiatry of Pandemics, 7–35 (2019), Springer, https://doi.org/10.1007/978-3-030-15346-5_2.

[2] Piret, J. and Boivin, G., Pandemics Throughout History. Front. Microbiol., 11:631736 (2021), doi: 10.3389/fmicb.2020.631736.

[3] Hays, J., Epidemics and pandemics: their impacts on human history (2005), ABC-CLIO, ISBN 978-1-85109-658-9.

[4] Porter, R., The Cambridge illustrated history of medicine (2001), Cambridge University Press, ISBN: 9780521002523.

[5] Stern, A.M. and Markel, H., The History Of Vaccines And Immunization: Familiar Patterns, New Challenges. Health Aff. 24–3 (2005), https://doi.org/10.1377/hlthaff.24.3.611.

[6] Plotkin, S.A. and Plotkin S.L., The development of vaccines: how the past led to the future, Nat Rev Microbiol. 9, 889–893(2011). https://doi.org/10.1038/nrmicro2668.

[7] Report of the WHO-China Joint Mission on Coronavirus Disease 2019 (COVID-19), World Health Organization (2019), https://www.who.int/docs/default-source/coronaviruse/who-china-joint-mission-on-covid-19-final-report.pdf, (Accessed on August 20, 2021).

[8] WHO Director-General’s opening remarks at the media briefing on COVID-19 - 11 March 2020, World Health Organization (2020), https://www.who.int/director-general/speeches/detail/who-director-general-s-opening-remarks-at-the-media-briefing-on-covid-19---11-march-2020, (Accessed on June 7, 2021).

[9] Worldometer, COVID-19 coronavirus pandemic, https://www.worldometers.info/coronavirus/, (Accessed on August 20, 2021)

[10] Jackson, J. et al., Global Economic Effects of COVID-19, Homeland Security Digital Library (2020), https://search.bvsalud.org/global-literature-on-novel-coronavirus-2019-ncov/resource/en/grc-740379, (Accessed on July 19, 2021).

[11] Donthu, N. and Gustafsson, A., Effects of COVID-19 on business and research, J Bus Res. 117: 284–289 (2020), https://doi.org/10.1016/j.jbusres.2020.06.008.

[12] Editorial Board, Research and higher education in the time of COVID-19, Lancet 396, 583 (2020), https://doi.org/10.1016/S0140-6736(20)31818-3.

[13] Pepita, B. et al., COVID-19 and the collapse of global trade: building an effective public health response, Lancet Planet. Health 5, 102–107 (2021), https://doi.org/10.1016/S2542-5196(20)30291-6.

[14] Tarricone, R. and Rognoni, C., What can health systems learn from COVID-19?, Eur Heart J Suppl. 22, 4–8 (2020), https://doi.org/10.1093/eurheartj/suaa185

[15] The pandemic has exacerbated existing political discontent, The Economist, https://www.economist.com/international/2021/07/31/the-pandemic-has-exacerbated-existing-political-discontent, (Accessed on July 19, 2021).

[16] Van Doremalen, N, et al., Aerosol and surface stability of SARS-CoV-2 as compared with SARS-CoV-1, N Engl J Med. 382, 1564–1567 (2020), doi.org/10.1056/NEJMc2004973.

[17] Tracking SARS-CoV-2 variants, World Health Organization (2021), https://www.who.int/en/activities/tracking-SARS-CoV-2-variants, (Accessed on July 16, 2021).

[18] The delta variant is the most dangerous SARS-CoV-2 mutation yet, The Economist (2021), https://www.economist.com/graphic-detail/2021/06/16/the-delta-variant-is-the-most-dangerous-sars-cov-2-mutation-yet, (Accessed on July 20, 2021).

[19] Fontanet, A. et al., SARS-CoV-2 variants and ending the COVID-19 pandemic, Lancet 397, 592–594 (2021), https://doi.org/10.1016/S0140-6736(21)00370-6

[20] British Society for immunology, Types of vaccines for COVID-19 (2021), https://www.immunology.org/coronavirus/connect-coronavirus-public-engagement-resources/types-vaccines-for-covid-19, (Accessed on Jun 29, 2021).

[21] Mathieu, E., Ritchie, H., Ortiz-Ospina, E. et al. A global database of COVID-19 vaccinations. Nat Hum Behav. 5, 947–953(2021). https://doi.org/10.1038/s41562-021-01122-8.

[22] Heida, R., et al., Inhaled vaccine delivery in the combat against respiratory viruses: a 2021 overview of recent developments and implications for COVID-19 (2021), Expert Rev. Vaccines, https://doi.org/10.1080/14760584.2021.1903878.

[23] Kim, J.H., Marks, F. & Clemens, J.D., Looking beyond COVID-19 vaccine phase 3 trials. Nat Med 27, 205–211 (2021). https://doi.org/10.1038/s41591-021-01230-y.

[24] Tang, Z. et al., Insights from nanotechnology in COVID-19 treatment, nanotoday 36, 101019 (2021), https://doi.org/10.1016/j.nantod.2020.101019.

[25] A new weapon in the war against SARS-CoV-2 has been found, The Economist (2021), https://www.economist.com/science-and-technology/2021/06/16/a-new-weapon-in-the-war-against-sars-cov-2-has-been-found. (Accessed on July 21, 2021).

[26] Mullard A., How COVID vaccines are being divided up around the world, Nature (2020), https://doi.org/10.1038/d41586-020-03370-6.

[27] America’s vaccination woes cannot be blamed only on politics, The Economist (2021), https://www.economist.com/united-states/2021/07/27/americas-vaccination-woes-cannot-be-blamed-only-on-politics. (Accessed on August 11, 2021).

[28] Brauer, F., Castillo-Chavez, C., & Feng, Z., Mathematical Models in Epidemiology, Texts in Applied Math., Springer (2019), New York, https://doi.org/10.1007/978-1-4939-9828-9.

[29] Brauer, F., Mathematical epidemiology: Past, present, and future, Infect Dis Model. 2-2, 113–127 (2017), https://doi.org/10.1016/j.idm.2017.02.001.

[30] Kermack, W. & Meckendrik A., A contribution to the mathematical theory of epidemics, Proc R Soc Lond. A115, 700–721 (1927), https://doi.org/10.1098/rspa.1927.0118.

[31] Brauer, F., The Kermack–McKendrick epidemic model revisited, Math Biosci. 198-2, 119–131 (2005), https://doi.org/10.1016/j.mbs.2005.07.006.

[32] Weiss, H., The SIR model and the Foundations of Public Health, Materials Matematics, 17 (2013), https://ddd.uab.cat/record/108432.

[33] Altmann, M., Susceptible-infected-removed epidemic models with dynamic partnerships, J. Math. Biology 33, 661–675 (1995). https://doi.org/10.1007/BF00298647.

[34] Fanelli, D. & Piazza, F., Analysis and forecast of COVID-19 spreading in China, Italy and France, Chaos Soliton Fract. 134, 109761 (2020), https://doi.org/10.1016/j.chaos.2020.109761.

[35] Baba, I.A., et al., Mathematical model to assess the imposition of lockdown during COVID-19 pandemic, Results Phys. 20, 103716 (2021), https://doi.org/10.1016/j.rinp.2020.103716.

[36] Viguerie, A., et al., Simulating the spread of COVID-19 via a spatially-resolved susceptible–exposed–infected–recovered–deceased (SEIRD) model with heterogeneous diffusion, Appl Math Lett. 111, 106617 (2021), https://doi.org/10.1016/j.aml.2020.106617.

[37] El Deeb, O. & Jalloul M., The dynamics of COVID-19 spread: evidence from Lebanon, Math Biosci Eng. 17-5, 5618–5632 (2020), https://doi.org/10.3934/mbe.2020302.

[38] Yang, C. & Yang J., A mathematical model for the novel coronavirus epidemic in Wuhan, China, Math Biosci Eng. 17-3, 2708–2724 (2020), https://doi.org/10.3934/mbe.2020148.

[39] Silva, P., et al., COVID-ABS: An agent-based model of COVID-19 epidemic to simulate health and economic effects of social distancing interventions, Chaos Soliton Fract. 139, 110088 (2020), https://doi.org/10.1016/j.chaos.2020.110088.

[40] Cuevas, E., An agent-based model to evaluate the COVID-19 transmission risks in facilities, Comput Biol Med. 121, 103827 (2020), https://doi.org/10.1016/j.compbiomed.2020.103827.

[41] Spassiani, I., et al., Spatiotemporal Analysis of COVID-19 Incidence Data, Viruses 13-3, 463 (2021), https://doi.org/10.3390/v13030463.

[42] El Deeb, O., Spatial Autocorrelation and the Dynamics of the Mean Center of COVID-19 Infections in Lebanon, Front Appl Math Stat. 6, 620064 (2021), https://doi.org/10.3389/fams.2020.620064.

[43] Gross, B., et al., Spatio-temporal propagation of COVID-19 pandemics, EPL 131, 58003 (2020), https://doi.org/10.1209/0295-5075/131/58003.

[44] Paltiel D., et al., Speed Versus Efficacy: Quantifying Potential Tradeoffs in COVID-19 Vaccine Deployment, Ann Intern Med. 174-3, 568–570 (2021), https://doi.org/10.7326/M20-7866.

[45] Chagla, Z., The BNT162b2 (BioNTech/Pfizer) vaccine had 95% efficacy against COVID-19 ≥7 days after the 2nd dose, Ann Intern Med. 174-2, JC15 (2021), https://doi.org/10.7326/ACPJ202102160-015.

[46] Knoll, M.D. and Wonodi, C., Oxford–AstraZeneca COVID-19 vaccine efficacy, Lancet 397, 72–74 (2021), https://doi.org/10.1016/S0140-6736(20)32623-4.

[47] Baraniuk, C., What do we know about China’s covid-19 vaccines?, BMJ 373, n912 (2021), https://doi.org/10.1136/bmj.n912.

[48] Covid: Brazil approves and rolls out AstraZeneca and Sinovac vaccines, BBC (2021), https://www.bbc.com/news/world-latin-america-55699535, (Accessed on August 1, 2021).

[49] Van Tulleken, C., Covid-19: Sputnik vaccine rockets, thanks to Lancet boost, BMJ 373, n1108 (2021), https://doi.org/10.1136/bmj.n1108.

[50] Logunov D.Y., et al., Safety and immunogenicity of an rAd26 and rAd5 vector-based heterologous prime-boost COVID-19 vaccine in two formulations: two open, non-randomised phase 1/2 studies from Russia, Lancet 396, 887–897 (2020), https://doi.org/10.1016/S0140-6736(20)31866-3.

[51] Chagla, Z., In high-risk adults, the Moderna vaccine had 94% efficacy against COVID-19 ≥14 d after the 2nd dose, Ann Intern Med. 174-3, JC28 (2021), https://doi.org/10.7326/ACPJ202103160-028.

[52] Center for Disease Control and Prevention (CDC), Interim Guidance on Ending Isolation and Precautions for Adults with COVID-19 (2021), https://www.cdc.gov/coronavirus/2019-ncov/hcp/duration-isolation.html. (Accessed on June 29, 2021).

[53] Lacobucci, G., Covid-19: Single dose of Pfizer and Oxford vaccines cuts risk of hospital admission by 80% in over 80s, data suggest, BMJ 372, n612 (2021), https://doi.org/10.1136/bmj.n612.

[54] Bernal, J.., et al., Effectiveness of the Pfizer-BioNTech and Oxford-AstraZeneca vaccines on covid-19 related symptoms, hospital admissions, and mortality in older adults in England: test negative case-control study, BMJ 373, n1088 (2021), https://doi.org/10.1136/bmj.n1088.

[55] Moghadas, S., et al., The Impact of Vaccination on Coronavirus Disease 2019 (COVID-19) Outbreaks in the United States, Clin Infect Dis. ciab079 (2021), https://doi.org/10.1093/cid/ciab079.

[56] Sette, A. & Crotty S., Adaptive immunity to SARS-CoV-2 and COVID-19, Cell 184-4, 861–880 (2021), https://doi.org/10.1016/j.cell.2021.01.007.

[57] Vinceti, M., et al., SARS-CoV-2 infection incidence during the first and second COVID-19 waves in Italy, Environ Res. 197, 11097 (2021), https://doi.org/10.1016/j.envres.2021.111097.

[58] Mazzaferro, S., et al., Waves of infection and waves of communication: the importance of sharing in the era of Covid-19, J Nephrol. 34, 633–636 (2021), https://doi.org/10.1007/s40620-021-00974-7.

